# Mediating role of Interleukin-6 in the predictive association of diabetes with Hippocampus atrophy, Amyloid, Tau, and Neurofilament pathology at pre-clinical stages of diabetes-related cognitive impairment

**DOI:** 10.1101/2025.05.06.25327092

**Authors:** Asma Hallab, The Health and Aging Brain Study (HABS-HD) Study Team

## Abstract

**Introduction:** Type-2 diabetes (T_2_DM) has been associated with higher dementia risks, but the mechanisms are still unclear, and there is increasing evidence of the role of cytokines. Interleukin-6 (IL-6) mediating effect has never been explored.

**Methods:** The study included a subset of 1,927 participants from the Health and Aging Brain Study: Healthy Disparities (HABS-HD) cohort with complete data. Cross-sectional and longitudinal analyses were performed. Associations were studied using multivariable linear, logistic, and mediation analysis with non-parametric bootstrapping.

**Results:** T_2_DM and IL-6 were associated with worse executive function, Hippocampus atrophy, lower Aß_42_/Aß_40_ ratio, and higher Aß_40_, Aß_42_, total Tau, and NfL levels. IL-6 mediated 5% of the association of T_2_DM with Aß_40_ ([1.5%-10%], *p-*value<2×10^−16^), 4% with Aß_42_ ([0.7%-11%], *p-* value=0.014), 8% with TMT-B ([0.2%-35%], *p-*value=0.046), 11% with total Tau ([2.5%-40%], *p-*value=0.010), 5% with NfL ([1.6%-8%], *p-*value<2×10^−16^), and 12% hippocampus atrophy ([3%-49%], *p-*value=0.004). The results, except TMT-B, were replicated in the longitudinal analysis of long-lasting T_2_DM on non-previously diagnosed cognitive impairment.

**Conclusions:** The study captured a pre-clinical stage of the T_2_DM-dementia association. The mediating effect of IL-6 is a novelty that has to be further explored and accounted for in risk stratification and preventive measures, particularly in ethnic minorities.

**Graphical abstract:** 

## 1. Introduction

Type 2 Diabetes Mellitus (T_2_DM) is one of the most geographically and demographically widespread non-communicable diseases (1) and is recognized as a global public health challenge owing to its association with increased morbidity and mortality rates. (2) Despite treatment availability, recent studies have raised serious concerns about the rapidly rising incidence (3) and insufficient coverage of specialized medical care, particularly among people with disadvantaged backgrounds and in lower-resource settings. (1) The asymptomatic evolution of diabetic pathophysiology over the first years leads to its discovery at advanced stages when “classical” and non-classical complications have already evolved. (4) While some of them might be reversible with adherence to regular treatment and healthy lifestyles, others might cause severe organ-specific damage and chronic disability. (5)

Cognitive disorders and dementia have also recorded increasing trends in the last decades, primarily owing to the growth and aging of the world population, (6) but also due to the changing lifestyles, and emerging obesity and glucose-related disorders in both industrialized and developing regions. (7) It is estimated that around 153 million persons will be affected by dementia in 2050. (6) Alzheimer’s disease (AD) is characterized by Amyloid, Tau, and Neurofilament chain (NfL: a newer biomarker of neurodegeneration) pathology (ATN), which summarizes neural injury and accumulation of amyloid and tau plaques in the brain. It is the most prevalent dementia type worldwide, followed by vascular dementia. (8) While each of these dementia types has a distinct pathophysiology, (9) they generally tend to co-occur in older adults. (10)

T_2_DM demonstrates complex and multifaceted associations with neuropsychiatric and cognitive disorders. (11–13) Microvascular complications are well-recognized mechanisms described in stroke (deep hemorrhagic stroke and lacunar ischemic stroke) and consequent dementia in patients with T_2_DM. (14) Thus, the association of T_2_DM with non-vascular brain pathology, (15) low-grade inflammation, (16) and the multisystemic aspect of immunity-related complications (17, 18) implies the involvement of other actors that need to be better understood. Insulin resistance is a key mechanism in T_2_DM, causing impaired glucose metabolism in different organs, including the brain and, specifically, neurons, glia, and astrocytes, all of which are provided with insulin receptors. (19) Insulin receptors are also located in the blood-brain barrier, and their impairment is associated with AD pathology. (20) On the other side, aging, social stress, overnutrition, disruption of the circadian rhythm, and sedentary lifestyles can trigger brain insulin resistance, which results in an increase in sympathetic nerve activity and reduces the activity of the Vagus nerve. (19) This leads, respectively, to lipotoxicity and hyperglycemia and, consequently, to systemic insulin resistance. (19) Brain insulin resistance is also associated with impaired body weight regulation and increased visceral fat distribution, increasing the risk of cardiometabolic complications. (21, 22) High plasma lipid and glucose levels are associated with elevated systemic pro-inflammatory cytokines and chronic inflammatory remodeling in various tissues. (23) This status of low-grade inflammation predicts the onset and evolution of several neuropsychiatric disorders. (24–26) Low-grade inflammation might, therefore, play a significant role in diabetes-related neurodegeneration and cognitive decline. While the evidence of reciprocal brain-insulin-body crosstalk has been largely discussed in the last few years, the concomitant role of inflammation remains unclear.

Understanding the potential implications of systemic inflammation in the association between T_2_DM and cognitive impairment in middle-aged and older adults has promising potential to provide new pathways toward preventive and therapeutic options. However, most published studies explored simplistic associations in cross-sectional designs and were limited to two of those three variables in each of their study question. None explored the integrated role of Interleukin-6 (IL-6) in the diabetes-brain constellation, specifically whether there is a mediating effect of IL-6 on the path between diabetes and biomarkers of cognitive decline. Furthermore, previous studies were performed in middle-aged and predominantly White populations, and there is a lack of data on older adults and less-represented ethnic groups, those known for being exposed to higher risks of insulin resistance and cognitive decline. (3, 27)

The first aim of this study was to assess the mediating effect of IL-6 in the association between T_2_DM and biomarkers of neurodegeneration and cognitive impairment. The second aim was to evaluate the mediating role of IL-6 in the longitudinal association between long-lasting T_2_DM and neurodegeneration-specific biomarkers in community-dwelling middle-aged and older adults without previously diagnosed cognitive decline.

## 2. Methods

The study has been performed following the Strengthening the Reporting of Observational Studies in Epidemiology (STROBE) guidelines. (28)

### 2.1. Study population

The study population is a subset of the Health and Aging Brain Study (HABS-HD) cohort. This NIH-funded study was performed at the Institute for Translational Research - University of North Texas Health Science Center as an extension of the HABLE study. (29) The first phase was initiated in 2017 with the general aim of studying health outcomes in Mexican Americans compared to non-Hispanic White controls. From 2021, 1,000 Black participants were enrolled in the cohort. HABS-HD is a community-based study; participants were recruited at several social events, and the study has been advertised in the media. Participants were encouraged to further advertise for the study, and snowball enrollment was commonly reported. Adults aged 50 and older were included, and those with type 1 diabetes, any form of dementia other than Alzheimer’s type, severe mental illness (except depression and anxiety), alcohol and substance use disorders, active infection or cancer, and severe physical illnesses interfering with cognition (exp., end-stage chronic kidney disease, chronic heart failure) were not eligible. (29)

Recruited participants underwent clinical, neuropsychiatric, biological, and neuroimaging investigations at baseline and a follow-up rhythm of 24-30 months intervals. However, many participants were lost to follow-up at 24 months, and, therefore, the current analysis is based on the baseline data of the 6^th^ data release.

Procedures contributing to this work were all compliant with the [Helsinki Declaration](https://www.wma.net/policies-post/wma-declaration-of-helsinki-ethical-principles-for-medical-research-involving-human-subjects/) of 1975, as revised in 2013, and with the ethical standards of the relevant national and institutional committees on human experimentation. Ethical approval was obtained from the local institutional review board (University of North Texas Health Science Center Institutional Review Board). Written informed consent was obtained from every participant. The current research consists of a secondary analysis of anonymized data and was performed in accordance with the data use agreement, followed by an authorization for publication.

### 2.2. Exposure

The diagnosis of T_2_DM was based on self-disclosure of medical history of T_2_DM, fasting blood glucose (mg/dL) levels, glycated hemoglobin (HbA1c) (%), type of medication (self-disclosed), and date of onset (self-disclosed). As previously mentioned, type 1 diabetes was an exclusion criterion from the source cohort. No clinical recorders were consulted in this community-based study.

Other T_2_DM-relevant measures were reported to describe the population: fasting serum Glucose levels (mg/dL), homeostatic model assessment for insulin resistance (HOMA-IR), age of onset of T_2_DM, type of treatment, healthy diet, and regular physical activity.

### 2.3. Outcomes

### 2.3.1. Cognitive impairment

The main outcome of the study was cognitive impairment as a binary variable at baseline. “No cognitive impairment” was the reference negative value (Ref=0). The presence of mild cognitive impairment (MCI) or dementia was labeled as “cognitive impairment” and was accorded a positive value (positive=1). Questions on past medical history were analyzed to identify cases with no priorly diagnosed cognitive impairment or dementia (any etiology).

Furthermore, results of the mini-mental status examination (MMSE) total score (in points) (30) and trail-making-test B (TMT-B) (in seconds) (31) were reported and evaluated.

#### 2.3.2. Amyloid ß_42_ and ß_40_, total Tau, phosphorylated Tau_181_, and NfL

Amyloid ß_42_ (Aß_42_) and ß_40_ (Aß_40_) levels were assessed in plasma and reported in pg/mL. A ratio was calculated based on the value of Aß_42_ on Aß_40_ levels. Similarly, total Tau, phosphorylated (p-Tau_181_), and Neurofilament Chain (NfL) levels were measured in plasma and reported in pg/mL. To ensure the homogeneity of the findings, only values that were assessed during the same phase using the Quanterix ultra-sensitive Simoa® (Single Molecule Array) technology were considered.

#### 2.3.3. Hippocampus volume

All participants underwent 3 Tesla magnetic resonance imaging (MRI) at the same site (two RMI scanners were used: MAGNETOM Skyra and MAGNETOM Vida 1, Siemens Healthineers^®^) and based on the ADNI 3 protocol. (32) T1-based segmentation was performed using Hippodeep. The Hippocampus volume was calculated as the sum of the right and left Hippocampi and reported in mm^3^. Hippodeep-measured intracranial volume (ICV) (mm^3^) and the version of MRI scanners were also reported and adjusted for.

### 2.4. Mediator

IL-6 was measured in the fasting blood plasma at baseline and reported in pg/mL. It was also assessed using the Quanterix ultra-sensitive Simoa® (Single Molecule Array) technology at the same study phase with plasma ATN measurements.

### 2.5. Confounders

Age (years), sex (females vs. males), ethnicity (self-disclosed “non-Hispanic White”, “Hispanic”, and “Black”), educational level (years), Apolipoprotein E (APOE) ε4 positivity (negative when none, and positive when one or two ε4 alleles are present), current tobacco smoking (binary), alcohol consumption (binary), and body-mass index (BMI); calculated as weight (Kg)/Height (m)^2^.

Models where Hippocampus volume was included as the dependent variable were additionally adjusted to the intracranial volume (ICV) (mm^3^) and MRI Scanner type. Models where Aß_40_, Aß_42_, their ratio, total Tau, p-Tau_181_, and NfL were included as dependent variables were additionally adjusted to the estimated glomerular filtration rate as a continuous variable (mL/min/1.73 m^2^), calculated based on the race-free CKD-EPI_2021_ equation. (33)

### 2.6. Inclusion criteria

Only cases with complete data on IL-6, Aß_40_, Aß_42_, total Tau, p-Tau_181_, NfL, and total Hippocampus volume were included. All cases had fully available information on diabetes and cognition-related diagnoses. Cases missing the MMSE total score or TMT-B duration were eligible.

### 2.7. Statistical analysis

The statistical analysis was performed with RStudio version 2024.12.1 (Posit Software^®^, PBC, USA). For the selection of relevant outcomes, false-discovery rate (FDR) adjusted *p-*values were calculated to reduce the risk of Type I error. The significance level of the two-sided *p-* or *p_FDR_-*values was set at 0.05.

#### 2.7.1. Dealing with missing values in covariates

Covariates’ values missing completely at random were counted and reported as proportions (%). When the missing proportion was <1%, continuous variables were imputed by the corresponding median value, and count variables by the null value.

#### 2.7.2. Exploring the normality of the distribution

Raw variables and their log-transformed values were plotted separately, and plots were overlaid with their corresponding kernel density curve (red), as well as a predicted curve for normal distribution (green), to evaluate the skewness in the data. The Shapiro-Wilk test was performed for each, and when the W-value increased through log transformation, this new log-scaled variable was used for the regression models and mediation analysis. Normally distributed variables, which satisfied the linearity assumption, were kept in their raw form since each outcome was explored independently. Largely skewed variables where the log transformation did not improve their distribution were dichotomized into clinically relevant categories.

#### 2.7.3. Data description and group comparison

Percentages (%) and median with interquartile range (IQR) were used to present data. Raw variables were included at this stage. Groups were compared using the Wilcoxon rank sum test or Kruskal-Wallis rank sum test for continuous variables and Pearson’s *Chi*-squared test (including Fisher’s exact test) for count variables. Associations were explored using adjusted regression linear models for continuous outcome variables and adjusted logistic regression models when the outcome of interest was binary. Interaction terms were introduced when indicated.

#### 2.7.4. Cross-sectional analysis

To understand the statistical relationship between different variables, confounder-adjusted regression analysis was based on T_2_DM or IL-6 levels (pg/mL) as an independent variable, cognition-specific biomarkers as binary (cognitive impairment, MMSE-impairment, TMT-B impairment), or as continuous dependent variables (plasma Aß_40_ & Aß_42_ levels (pg/mL), their corresponding ratio (Aß_42_/Aß_40_), Tau and p-Tau_181_ levels (pg/mL), NfL (pg/mL), and total Hippocampus volume (mm^3^)). Corresponding *p-*values were adjusted for FDR. Only associations with *p_FDR_*-value <0.05 (in both T_2_DM and IL-6 corresponding models) were eligible for mediation analysis.

#### 2.7.5. Mediation analysis

Mediation analysis with 1000-fold non-parametric bootstrapping of 95% CI based on the percentile method was applied. The same confounders were introduced in the mediator and outcome models. Hippocampus-specific mediation analysis was additionally adjusted for ICV (mm^3^) and type of MRI scanner. Plasma ATN-specific mediation analysis was additionally adjusted for eGFR value (mL/min/1.73 m^2^). The estimates of the mediated effect (ACME), direct effect (ADE), and total effect were visualized with their corresponding 95% CI. Rounding to the second decimal was applied, except for variables with very low effect values. The proportion mediated was calculated as the value of the mediated effect from the total effect and reported in a fully rounded percentage (for an easier interpretation of the data) with the corresponding 95% CI and *p-*value.

#### 2.7.6. Longitudinal analysis

To understand the underlying pathophysiology in a longitudinal framework, a second analysis was performed. After excluding cases with prior cognitive impairment or dementia diagnosis and those with recently diagnosed diabetes (within the last five years), mediation analysis was based on long-lasting T_2_DM (at least for five years) as exposure, FDR-adjusted cognition-specific biomarkers as an outcome (baseline data), and IL-6 (pg/mL) (baseline data) as mediator. The same steps were followed for the mediation analysis.

## 3. Results

### 3.1. Included data

The main analysis included a total of 1,927 participants with complete data.

Missing variables affected BMI data (n=12, 0.6%), APOE ε4 status (n=7, 0.4%), and creatinine/eGFR (n=19, 0.9%). BMI and eGFR were imputed by median values, while missing APOE ε4 values were imputed by a null value.

Plasma Aß_40_ (pg/mL), amyloid ratio (Aß_42_/Aß_40_), Tau, p-Tau_181_ (pg/mL), NfL (pg/mL), plasma Glucose (mg/dL), HbA1c (%), HOMA-IR, and IL-6 (pg/mL) levels were strongly right skewed, therefore, log-transformed for the linear regression and mediation analyses. Plasma Aß_42_ (pg/mL) and total Hippocampus volume (mm^3^) had a normal distribution and were kept in their raw form (otherwise distorted by log-transformation). MMSE and TMT-B total scores were converted to categorical data based on relevant cutoff values (24 points for MMSE and 90 seconds for TMT-B).

### 3.2. Study population

Around one-quarter of the included cases were diagnosed with T_2_DM (24.13%). The median age was 66 years, and 62% were females. For age and sex, no statistically significant differences were observed between cases with and without T_2_DM. Details on the total population and group comparison are provided in **Table 1**.

T_2_DM was significantly more prevalent among Hispanics (65% vs. 23% White and 12% Black), while more White participants did not have T_2_DM (51% vs. 36% Hispanic and 12% Black, *p*-value<0.001). Participants with T_2_DM tend to be, on average, two years less educated than those without diabetes (12 vs. 14 years, *p*-value<0.001). Biologically, participants with T_2_DM had higher plasma levels of fasting Glucose, HbA1c, HOMA-IR, and IL-6 in addition to higher BMI. While the rate of alcohol consumption did not significantly differ between groups, participants with T_2_DM disclosed significantly higher Tobacco smoking (8% vs. 5.1%, *p*-value=0.023).

Depression (41% vs. 32%, *p-*value<0.001), anxiety (21% vs. 16%, *p-*value=0.030), hypertension (79% vs. 59%, *p*-value<0.001), obesity (57% vs. 42%, *p-*value<0.001), and dyslipidemia (79% vs. 67%, *p-*value<0.001) were significantly more prevalent in T_2_DM participants. All cognitive biomarkers, except plasma p-Tau_181_ levels, were significantly worse in the T_2_DM group (all *p*-values<0.001, except p-Tau_181_ not significant).

Remarkably, a lower number of participants with APOE ε4 alleles had T_2_DM (21% vs. 27%, *p-* value=0.006). Similarly, although a higher number of participants with T_2_DM disclosed having chronic kidney dysfunction (6.2% vs. 2.9%, *p*-value=0.001), eGFR was significantly higher in this T_2_DM group (91 vs. 86 mL/min/1.73 m^2^, *p*-value=0.003).

### 3.4. Diabetes characteristics and group comparisons

Of the 465 total cases with diabetes at baseline, most of them (n=399 to 416) reported details on diabetes-related habits. The majority of participants with diabetes are under any type of treatment (88%), following healthy eating habits (83%), and keeping regular physical activity (64%). Hispanic and Black participants disclosed younger ages of T_2_DM onset than White participants (50 vs. 59 years in White, *p*-value<0.001). Cognition and ethnicity-based group comparison are detailed in **Table 2**.

### 3.5. IL-6 levels and associations with metabolic biomarkers

Higher IL-6 levels were significantly associated with higher Glucose (log-scaled adj. *ß*=0.04 [0.02, 0.06], *p-*value<0.001), HbA1c (log-scaled adj. *ß*=0.03 [0.02, 0.04], *p-*value<0.001), and HOMA-IR (log-scaled adj. *ß*=0.19 [0.10, 0.27], *p-*value<0.001) values. Similarly, higher glucose (log-scaled adj. *ß*=0.23 [0.12, 0.34], *p-*value<0.001), HbA1c (log-scaled adj. *ß*=0.038 [0.21, 0.54], *p-*value<0.001), and HOMA-IR (log-scaled adj. *ß*=0.09 [0.05, 0.13], *p-*value<0.001) levels were associated with increased IL-6 levels. Introducing an interaction term with diabetes showed significant effects. Interaction-specific visualization and stratification showed a dissociation in results, and the previous associations remained statistically significant only in non-T_2_DM participants. In T_2_DM, the associations lost their statistical significance. Results of the regression analysis are detailed and visualized in **Figure 1.a-f**.

**Figure 1:**
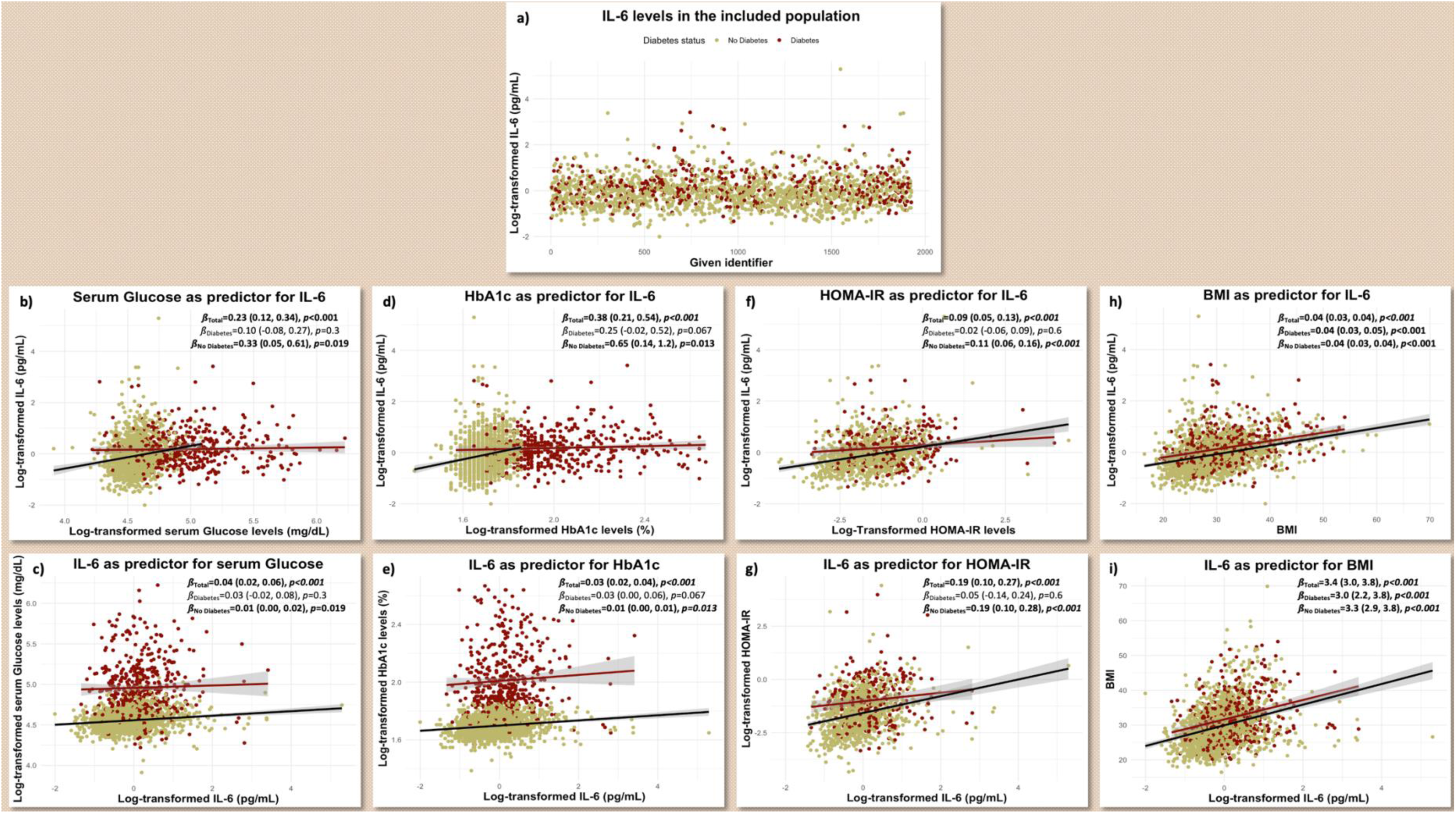
Interleukin-6 levels in the study population and associations with different metabolic biomarkers. **1.a)** Interleukin-6 (IL-6) in the study population. **1.b) & 1.c)** Serum Glucose levels. **1.d) & 1.e)** Glycated Hemoglobin (HbA1c). **1.f) & 1.g)** Homeostatic Model Assessment for Insulin Resistance (HOMA-IR). **1.h) & 1.i)** Body Mass Index (BMI). <u>Footnote</u>: Interactions between Interleukin-6 and diabetes are visualized.

The bilateral association between higher IL-6 levels and higher BMI remained statistically significant with the same coefficients in both diagnostic groups (**Figure 1.h & i**).

### 3.6. IL-6 and cognitive biomarkers

An increase in log-transformed IL-6 levels was significantly associated with 35% higher odds of cognitive impairment (OR=1.35 [1.13, 1.60], *p_FDR_-*value<0.001) and 29% higher odds of TMT-B ≥ 90 sec (OR=1.29 [1.07, 1.56], *p_FDR_-*value=0.011). It was also significantly associated with a decrease in the total Hippocampus volume (adj. *ß*=-92 [−140, −44], *p_FDR_-*value<0.001) and Aß_42_/Aß_40_ ratio (log-scaled adj. *ß*=-0.02 [−0.04, 0.00], *p_FDR_-*value=0.036), and an increase in the level of Aß_40_ (log-scaled adj. *ß*=0.04 [0.02, 0.06], *p_FDR_-*value<0.001), Aß_42_ (adj. *ß*=0.26 [0.08, 0.44], *p-*value=0.007), total tau (log-scaled adj. *ß*=0.05 [0.02, 0.07], *p_FDR_-*value<0.001), p-Tau_181_ (log-scaled adj. *ß*=0.03 [0.00, 0.06], *p_FDR_-*value=0.048), and NfL (log-scaled adj. *ß*=0.10 [0.07, 0.14], *p_FDR_-*value<0.001). No significant association was observed between IL-6 and the odds of MMSE ≤ 24 points. (**Table 3**)

No dissociation has been observed between T_2_DM and non-T_2_DM groups in this analysis (**Figure 2**).

**Figure 2:**
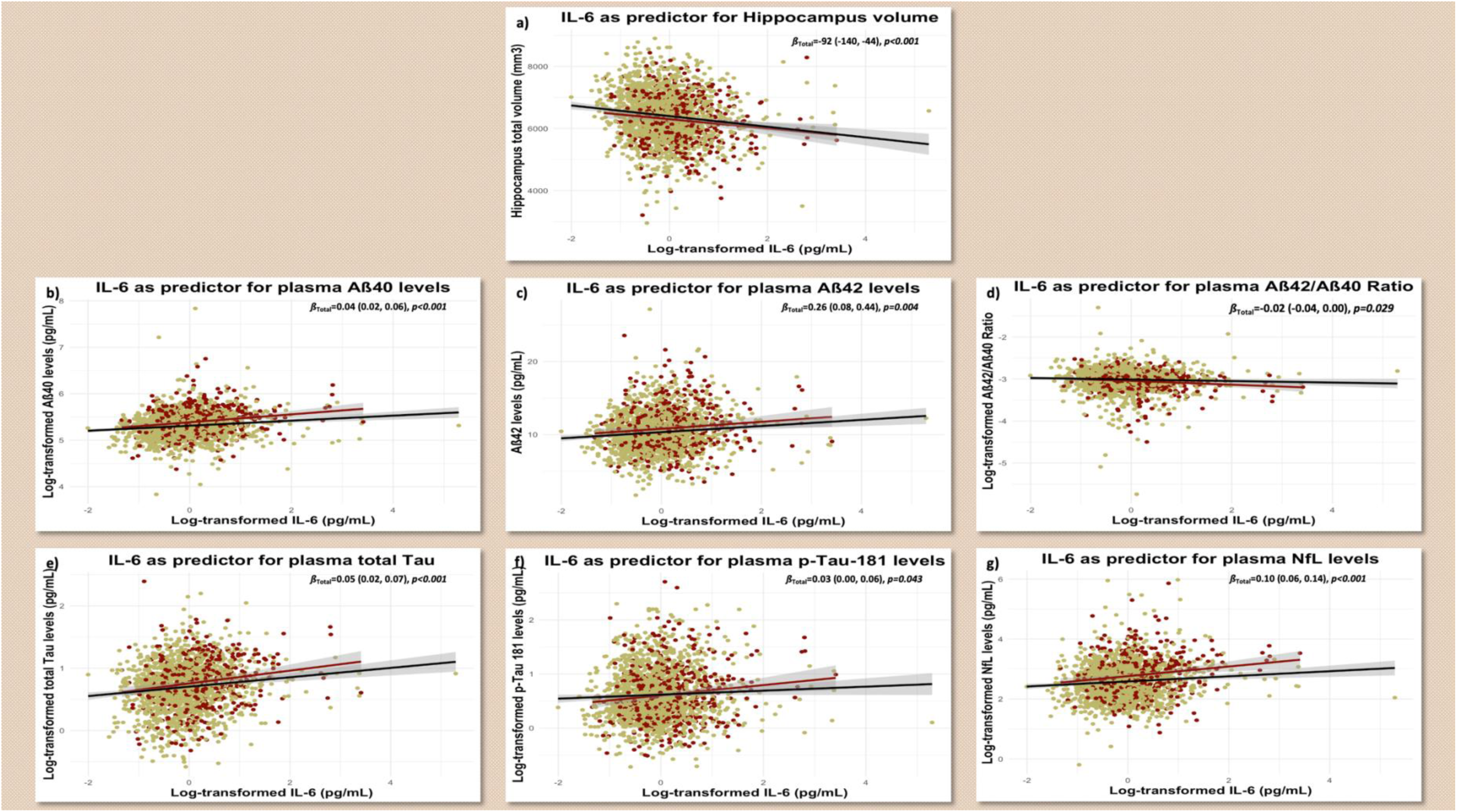
Association between Interleukin-6 levels and different cognitive biomarkers. Interactions between Interleukin-6 and diabetes are visualized. **2.a)** Hippocampus volume. **2.b)** Amyloid ß_40_. **2.c)** Amyloid ß_42_. **3.d)** Amyloid ß_42_/_40_ ratio. **4.e)** Total Tau. **2.f)** Phosphorylated (p-)Tau_181_. **3.g)** Neurofilament Chain. **<u>Footnote</u>**: Amyloid, Tau, and Neurofilament models are adjusted for renal function. Hippocampus models are adjusted for intracranial volume and scanner type.

### 3.7. Diabetes and cognitive biomarkers

Diabetes at baseline (independently from the duration of the disease) was significantly associated with 44% higher odds of TMT-B ≥ 90 sec (OR=1.44 [1.09, 1.90], *p_FDR_-*value=0.009), a decrease in total Hippocampus volume (adj. *ß*=-89 [−161, −17], *p_FDR_-*value=0.016) and Aß_42_/aß_40_ ratio (log-scaled adj. *ß*=-0.04 [−0.06, −0.01], *p_FDR_-*value=0.010), and an increase of Aß_40_ (log-scaled adj. *ß*=0.09 [0.07, 0.12], *p_FDR_-*value<0.001), Aß_42_ (adj. *ß*=0.63 [0.36, 0.89], *p_FDR_-* value<0.001), total Tau (log-scaled adj. *ß*=0.05 [0.01, 0.09], *p_FDR_-*value=0.012), and NfL levels (log-scaled adj. *ß*=0.24 [0.18, 0.29], *p_FDR_-*value<0.001).

There was also a significant association between having T_2_DM and higher IL-6 levels (log-scaled adj. *ß*=0.12 [0.05, 0.19], *p-*value<0.001).

There was no significant association between T_2_DM and the odds of cognitive impairment, MMSE total score ≤ 24 points, or p-Tau_181_ levels (after adjusting for multiple testing). (**Table 3**)

### 3.8. Mediation analysis: Cross-sectional design

The association between diabetes and biomarkers of cognitive impairment was significantly mediated by IL-6 (5% for Aß_40_ ([1.5%, 10%], *p-*value<2 × 10^−16^), 4% for Aß_42_ ([0.7%, 11%], *p-* value=0.014), 8% for pathological TMT-B ([0.2%, 35%], *p-*value=0.046), 11% for total Tau ([2.5%, 40%], *p-*value=0.010), 5% for NfL ([1.6%, 8%], *p-*value<2 × 10^−16^), and 12% for Hippocampus atrophy ([3%, 49%], *p-*value=0.004)). IL-6 did not show a significant mediating role in the association with the Aß_42_/Aß_40_ ratio (p-Tau_181_ was not included in the mediation analysis since it did not survive the FDR-adjusted associations). Statistically significant results are shown in **Figure 3**.

**Figure 3:**
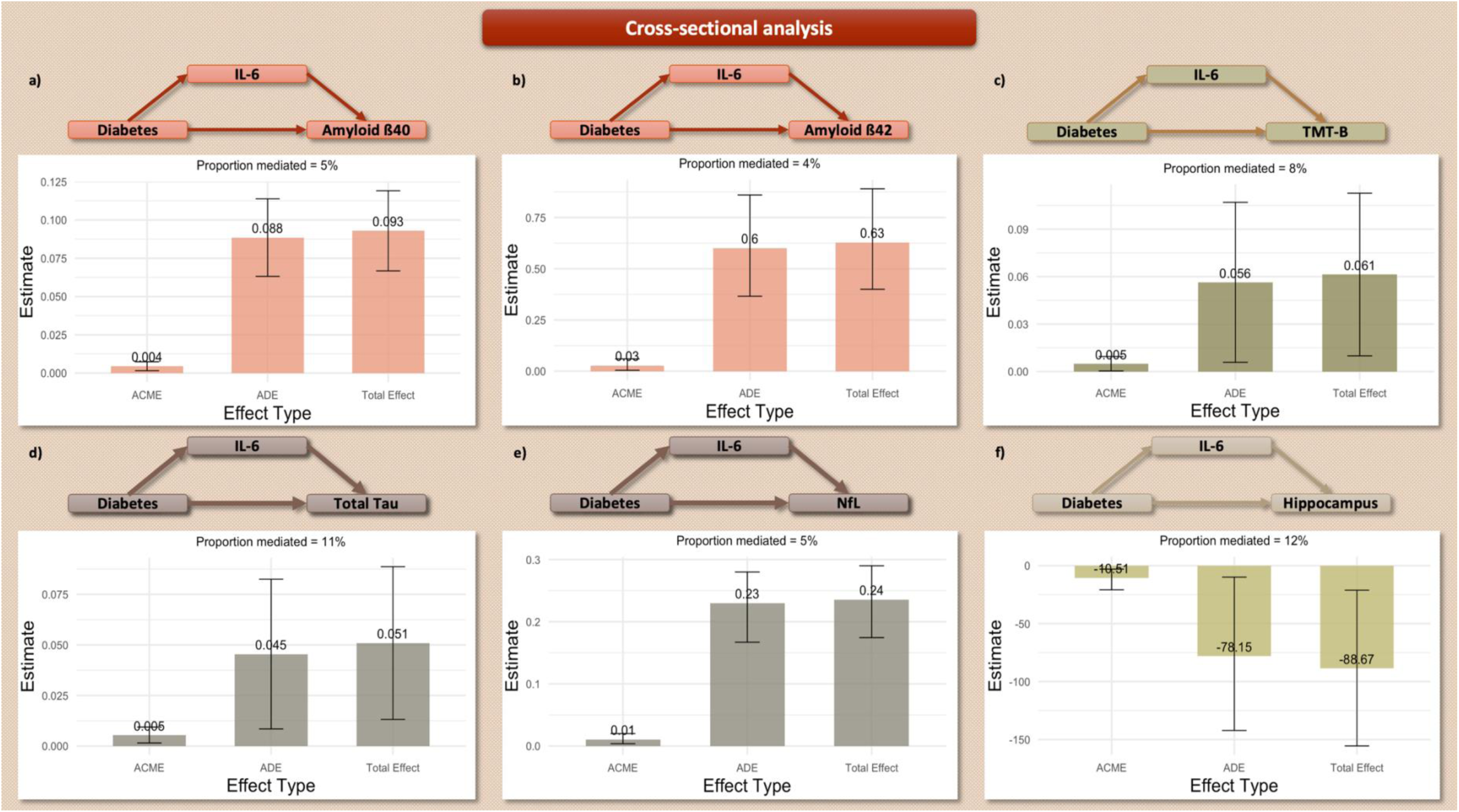
Mediation analysis – Cross-sectional design. Only statistically significant results are visualized. **3.a)** Amyloid ß_40_. **3.b)** Amyloid ß_42_. **3.c)** Trails-Making-Test B. **4.d)** Total Tau. **3.e)** Neurofilament Chain. **3.f)** Hippocampus volume. **<u>Footnote</u>**: ACME: Mediated Effect, ADE: Direct Effect. Amyloid, Tau, and Neurofilament models are adjusted for renal function. Hippocampus models are adjusted for intracranial volume and scanner type.

### 3.9. Mediation analysis: Retrospective longitudinal design

Particularities of this analyzed subgroup are analyzed and summarized in **Supplementary Tables 1 and 2**.

The association between long-lasting diabetes and biomarkers of cognitive impairment in cases without previously diagnosed cognitive impairment was significantly mediated by IL-6 (3% for Aß_40_ ([0.8%, 8%], *p-*value=0.008), 3% for Aß_42_ ([0.6%, 8%], *p-*value=0.014), 8% for total Tau ([1.6%, 41%], *p-*value=0.016), 3% for NfL ([0.7%, 6%], *p-*value=0.008), and 9% for Hippocampus atrophy ([1.5%, 37%], *p-*value=0.024)). Here also, IL-6 did not show a significant mediating role in the association with Aß_42_/Aß_40_ ratio (TMT-B and p-Tau_181_ were excluded from the mediation analysis). Statistically significant findings are visualized in **Figure 4**.

**Figure 4:**
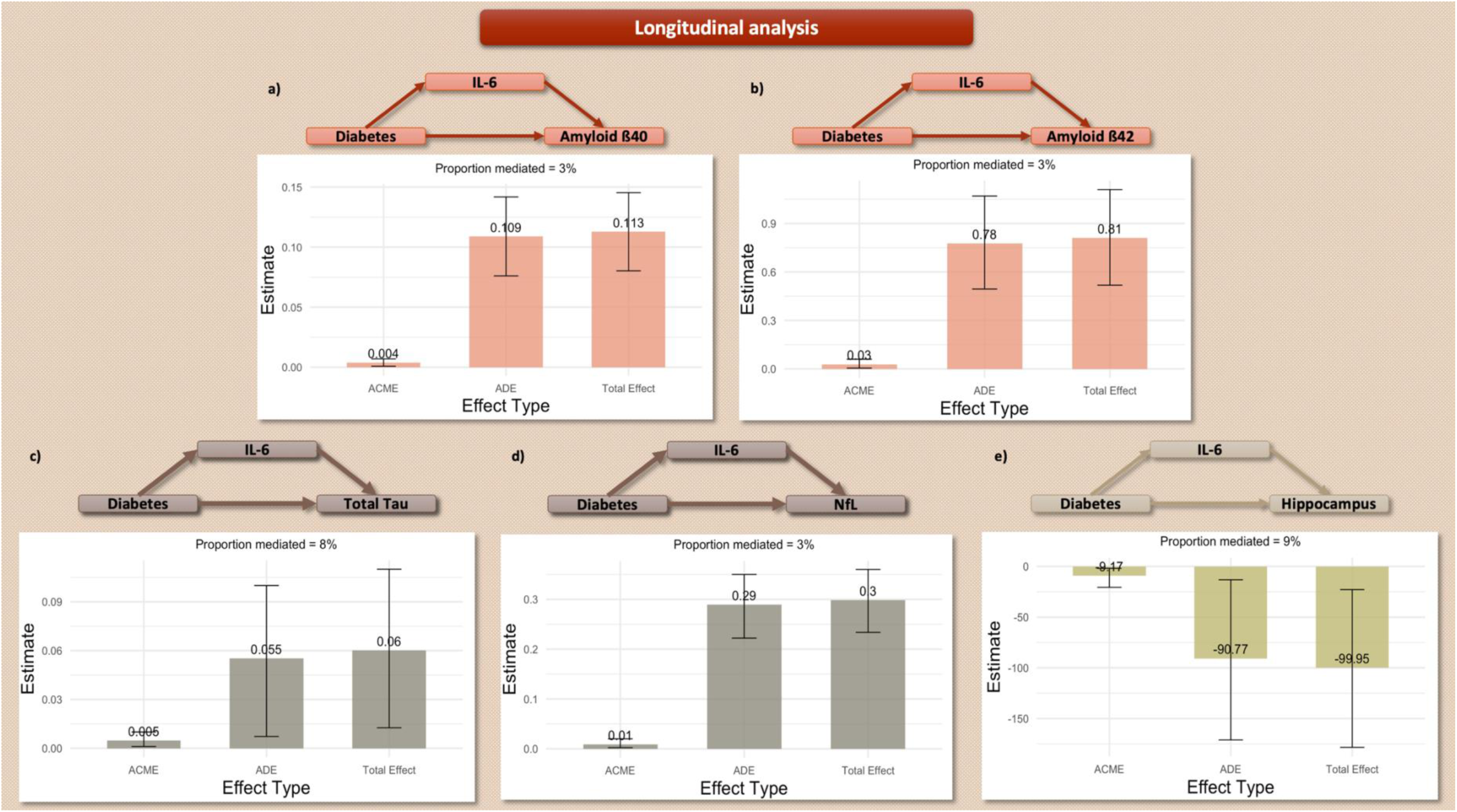
Mediation analysis – Longitudinal design. Only statistically significant results are visualized. **4.a)** Amyloid ß_40_. **4.b)** Amyloid ß_42_. **4.c)** Total Tau. **4.d)** Neurofilament Chain. **4.e)** Hippocampus volume. **<u>Footnote</u>**: ACME: Mediated Effect, ADE: Direct Effect. Amyloid, Tau, and Neurofilament models are adjusted for renal function. Hippocampus models are adjusted for intracranial volume and scanner type.

## 4. Discussion

The study showed significant associations between T_2_DM and several biomarkers of cognitive impairment, but not global cognition. IL-6 levels were higher among T_2_DM participants and mediated most observed associations. The results of the cross-sectional analyses were replicated in longitudinal analyses, where preexisting long-lasting T_2_DM had a significant predictive value on most cognitive biomarkers at baseline (particularly Hippocampus volume, total Tau, and NfL), despite excluding those with priorly diagnosed cognitive impairment. Several unexpected findings were revealed by this study. First, the lower proportion of APOE ε4 holders and higher eGFR levels in the T_2_DM participants. Second, the earlier onset age of T_2_DM in ethnic minorities. Finally, the T_2_DM-specific dissociation in the results of the association between IL-6 and diabetes-specific biomarkers (plasma Glucose, HbA1c, and HOMA-IR levels). Although non-primary, these findings can help understand the main results in their context.

### 4.1. Diabetes in older adults

A French multicenter study in older adults (11.1% with diabetes) showed that more adiposity (32.5 vs. 11.2%, *p-*value<0.001), hypertension (77.4 vs. 59.8%, *p-*value<0.001), dyslipidemia (60.5 vs. 48.9%, *p-*value=0.002), and cardiovascular history (19.3 vs. 7.7%, *p-*value<0.001) were found in those with diabetes. The cognitive MMSE test scores were significantly worse in cases with diabetes than in those without. These results are comparable to our cohort. However, the prevalence of T_2_DM and obesity were much higher in our US-based cohort.

As expected, more participants with chronic kidney disease were among those who had diabetes (cross-sectional and longitudinal analyses). The unexpected finding was, however, the higher eGFR value in the T_2_DM group compared to the healthy control. For the current analysis, the eGFR was calculated based on the race-free CKD-EPI_2021_ formula. The results were verified using the CKD-EPI_2009_ formula-based eGFR (non-showed data), and the findings were replicated. The discrepancy might be explained by the early-stage diabetes-related hyperfiltration, where affected persons exhibit supraphysiological higher renal filtration rates in order to hyper-compensate for nephron damage. (34) Glomerular hyperfiltration is also a biomarker of early kidney function impairment in pre-diabetes and pre-hypertension (35) and predicts higher mortality risk. (36) In the longitudinal subset of the study, where only long-lasting diabetes was considered, although higher rates of chronic kidney disease were found in the T_2_DM group, no significant difference in eGFR values between the groups was observed. Other hypotheses might include the protective effect of medications, (34) eating habits, (37) and selection bias. This bias might be induced by the inclusion of more health-seeking and compliant participants in the diabetes groups, in addition to the low number of those with chronic kidney disease (only those at very early stages). This theory might be validated by the restrictiveness of the inclusion criteria, as participants with severe chronic kidney disease were not eligible for HABS-HD. This potential bias is, however, a strength in the estimation of the plasma ATN levels, since their levels might be significantly influenced by the renal function. (38) All our plasma-based ATN-specific regression models (including mediation analysis) were adjusted for eGFR, which had a significant effect on the corresponding models, but did not change the significance level of the observed results.

Like in our current cohort, the French study reported fewer APOE ε4 carriers in the diabetes group than in controls (24.6 vs. 30.6%, *p-*value=0.05). (39) Previous studies found, however, an increased risk of insulin resistance in APOE (ε4/ε4) persons, exercising a synergetic interaction on the alteration of brain-blood barrier integrity. (40) A hypothetical explanation for these discrepancies might be associated with an earlier onset of AD, diabetes, and related complications in APOE ε4/ε4 persons, and, therefore, the lower likelihood of their participation in this cohort of community-dwelling adults. Furthermore, the mortality risk is higher in those with an earlier onset of T2DM. (2, 41) Ethnic minorities, whose age of onset of T_2_DM was earlier than White counterparts, need particular attention, as suggested by our data.

### 4.2. Diabetes and cognition

Cross-sectional studies using UK-Biobank data showed significant associations between diabetes and specific cognitive domains (intelligence, numeric memory, reaction time…), while one longitudinal study found a significant association between (pre-)diabetes and increased risk of cognitive decline. (11) The French cohort study found that neurodegeneration, but not small vessel disease or AD pathology, mediated the association between diabetes and worse cognition. (39) The authors evaluated neurodegeneration through the mean right and left cortical thickness, brain parenchymal fraction, mean glucose uptake at positron emission tomography (PET) scans, and total Hippocampus volume. In contrast, our current analysis considered the Hippocampus volume as an outcome, not a mediator. Hippocampus volume is a sensitive biomarker for early neurodegenerative processes and precedes other structures in predicting cognitive decline. (42) Furthermore, the French cohort found no mediating effect of the Aß_42_/Aß_40_ ratio, total Tau, and p-Tau between diabetes and cognitive decline. (39) While they used cerebrospinal fluid (CSF)-based values, our current study included plasma-based measurements. CSF levels might be more sensitive than plasma levels. However, our choice is primarily due to data availability, and our analysis of plasma ATN levels was based on highly sensitive technologies enabling high accuracy of cognitive screenings in community-based settings. (43) In our study, T_2_DM predicted a significant decrease in the plasma-based Aß_42_/Aß_40_ ratio, reflecting an association with a sensitive biomarker of AD pathology. Similarly, our study found a significant association between T_2_DM and higher plasma total Tau and p-Tau_181_ levels, contradicting the French cohort findings, which found a significant association between diabetes and higher Amyloid load only at PET imaging. A recent meta-analysis showed, however, that pooled results of different studies, including the latter, are not in favor of a significant association between diabetes and Amyloid pathology, neither in PET nor in CSF measures. (15) The pooled data from PET-and CSF-Tau studies, in contrast, showed a significant association of Tau pathology with Glucose metabolism disorders and diabetes combined. (15)

A recent study in older patients with T_2_DM and overweight or obesity explored longitudinal associations between cognition and plasma levels of Aß_40_, Aß_42_, total Tau, p-Tau_181_, glial fibrillary acidic protein (GFAP), and NfL. Only the increase over time in GFAP and NfL levels was a significant predictor of incident cognitive impairment and concomitant cognitive decline. (44) The selection bias related to the interventional intention of that study at baseline, the inclusion of high-risk participants with both T_2_DM and overweight and obesity, and the absence of a non-T_2_DM control group might explain the differences from our results. Furthermore, the included population was younger than 65 years at baseline, probably prior to the typical age of onset of detectable Amyloid pathology. Therefore, the study did not exclude the hypothesis of significant associations with Amyloid pathology in later ages. (44) The renal function had a significant impact on the predictive value of plasma-measured p-Tau_181_. (44) In our study, plasma ATN models were adjusted for renal function, and their statistical significance was not affected after correcting for eGFR. This implies that higher plasma ATN levels in T_2_DM are not explained by a reduced renal clearance.

The association between insulin resistance and AD has been largely discussed. Insulin resistance precedes AD pathology and is involved in the impairment of Aß clearance in the brain. (37) Plasma p-Tau is associated with Amyloid pathology and predicts cognitive decline. (45) The lack of significant associations in our study might indicate early stages of the AD continuum.

While the current study failed to find a significant association between T_2_DM and the overall state of cognitive impairment, another study showed that diabetes complications, mainly diabetic foot, microvascular, cerebrovascular, and cardiovascular diseases, were significant predictors of dementia onset within ten years of follow-up. (46) This might further suggest that our participants are at very early pre-clinical stages of diabetes-related complications.

Furthermore, insulin signaling disturbance in the brain is associated with impaired neurotransmitter signaling in preclinical models of psychosis. (47) This neurotransmitter hypothesis might be a further mechanism, but could not be explored by our cohort and needs further investigation.

### 4.3. Diabetes and IL-6

Higher IL-6 levels have a significant predictive value for incident diabetes. In a large, bi-ethnic US-based cohort, higher IL-6 predicted higher risks of incident diabetes and metabolic syndrome after 9.5 years of follow-up. (48) However, after stratification, the association was statistically significant only in non-Hispanic White participants, but not Black ones, despite higher IL-6 levels in the latter group at baseline. (48) This might be related to the ethnic discrepancy in the age of onset of diabetes in Black participants and survival-related selection bias. (49) This age gap was also demonstrated in our study, as Hispanic and Black participants with diabetes disclosed an earlier age of onset compared to White controls.

### 4.4. Cognition and IL-6

Several studies described the association between high IL-6 levels and cognitive impairment. A large UK-Biobank population study found a significant association between high IL-6 levels, cortical and subcortical atrophy (including the Hippocampus), and all-cause dementia in the longitudinal analysis. (50) The findings of the Berlin Aging Study showed a significant association between higher IL-6 levels and poorer executive function and processing speed in older adults (≥60 years). (51) In contrast, data from the Mayo Clinic Study of Aging did not show a significant association between IL-6 levels and domain-specific z-scores after adjusting for confounders. However, the latter study revealed a significant association between higher IL-6 levels and the concomitant odds of MCI in the cross-sectional analysis, but not incident cases during the follow-up. (52) IL-6 levels exhibit both between and within individual variations in the long term, which might reduce the specificity and statistical power of prospective predictive models.

### 4.5. T_2_DM, cognition, and IL-6

In the UK-based Edinburgh Type 2 Diabetes Study, higher baseline levels of IL-6 were significantly associated with a decline in abstract reasoning and executive function within a 10-year follow-up period. (53) IL-6 levels have not demonstrated a solid predictive value on global cognition in this UK study, (53) which confirms our current findings and highlights the domain-specific association between cognition, IL-6, and T_2_DM.

The difference between participants with and without T_2_DM in the statistical significance of association between higher IL-6 levels, and diabetes-related biomarkers (plasma Glucose, HbA1c, HOMA-IR) levels raises new questions on the eventual beneficial effect of diabetes-related therapy (medication, diet, regular physical activity) on down-regulating diabetes-related systemic inflammation. Previous interventional studies did not show a significant impact of insulin and metformin on the inflammatory biomarkers after a follow-up of 14 weeks, despite their positive effect in regulating blood glucose levels. (54) This does not exclude, however, the long-term effects of various therapeutic interventions (different diabetes-regulating medications, mental health support, stress reduction, (55) physical activity, (56) and Mediterranean diet (57)) on systemic inflammation in T_2_DM. These protective factors have also been associated with a decrease in dementia incidence. (58)

### 4.6. Summary

To summarize, the study showed a significant association between T_2_DM, executive function (only cross-sectional), Hippocampus atrophy, plasma Amyloid, Tau, and NfL levels. IL-6 mediated significantly most of these associations. These results, in addition to the absence of association with clinically relevant global cognitive impairment, might translate initial stages of the T_2_DM-dementia pathology, where kidney function and Amyloid production are in hyper-compensatory stages, despite the significant concomitant decrease of the Aß_42_/Aß_40_ ratio. Furthermore, despite the significant association with IL-6, p-Tau_181_ missed the statistical levels of significance in the association with T_2_DM and needs to be monitored in the longitudinal course. The dissociations observed between T_2_DM and non-T_2_DM groups in the IL-6-based associative analyses might indicate a beneficial effect of mediation, healthy diet, and physical activity, as most participants with T_2_DM seem to be compliant with at least one of these protective factors, and this hypothesis needs to be explored by larger interventional and registry/population-based studies.

### 4.7. Strengths

This is the first study based on two frameworks (cross-sectional and longitudinal) to assess the association between diabetes and biomarkers of AD and the mediating role of IL-6. The large number of included participants from various ethnic backgrounds, the large set and completeness of the collected data, and the monocentricity and homogeneity of the assessment methods are further strengths of this study. The use of a non-parametric bootstrapping method to estimate 95% CI in the mediation analysis presents a methodological novelty.

### 4.8. Limitations

The retrospective collection of medical history is the main limitation of the study. While it is less probable that there is a recall bias regarding the date of onset of diabetes, some participants, particularly those with cognitive impairment, might have provided imprecise dates. However, these cases were removed from the longitudinal analysis. Furthermore, diabetes might remain underdiagnosed for years, and many excluded participants from the longitudinal mediation analysis might have had diabetes, thus undiagnosed and first assessed during the recruitment in this cohort. This diagnosis bias might also have affected the assessment of cognitive impairment, as participants might have had a subtle form of cognitive impairment but remained undiagnosed until being recruited in the HABS-HD study. Another limitation is related to missing data on IL-6, Hippocampus volume, Amyloid, Tau, and NfL pathologies. Despite that, the number of recruited participants remained high compared to previous cohorts.

## 5. Conclusions

IL-6 mediates the association between long-lasting T_2_DM and AD biomarkers in middle-aged and older adults without previously diagnosed cognitive impairment. Despite the statistical significance of the observed associations, the mediating role of IL-6 does not explain the totality of the diabetes-related effect. This suggests other mechanisms besides IL-6-mediated neuroinflammation. More studies are needed to better understand further mediating risk factors, cluster patients based on associated predisposing factors, and orient toward personalized treatment.

## Declarations

### Ethical approval

All procedures contributing to this work comply with the ethical standards of the relevant national and institutional committees on human experimentation and with the Helsinki Declaration of 1975, as revised in 2013. Ethical approval was obtained from the local institutional review board (North Texas IRB Board). Participants gave written informed consent.

### Authorization for publication

The principal investigator and study director of the HABS-HD study revised the current version and ensured its compliance with DUA and authorized the publication of the manuscript.

### Authorship

AH has full access to all of the data and takes responsibility for the integrity of the data and the accuracy of the analysis, visualization, drafting, and editing of the manuscript.

### Data availability

Data can be acquired by qualified researchers after an official request.

## Acknowledgment

“Research reported on this publication was supported by the National Institute on Aging of the National Institutes of Health under Award Numbers R01AG054073, R01AG058533, R01AG070862, P41EB015922, and U19AG078109. The content is solely the responsibility of the authors and does not necessarily represent the official views of the National Institutes of Health.”

